# Quality of life and its associated factors among epileptic patients on treatment follow up in North Shoa administration, Amhara National State, Ethiopia

**DOI:** 10.1101/2022.12.28.22284016

**Authors:** Esubalew Guday, Getabalew Engdaye, Belachew Tekleyohannes, Nigus Alemnew, Akne Eshete, Yihenew Sewale

## Abstract

**Background:** Epilepsy is a common condition worldwide and has been observed to affect quality of life. Epilepsy patients have a lower quality of life than the general population as well as many other chronic disease patients. However, aside from focusing on symptom reduction, no attention is paid to the quality of life of those with epilepsy. This study aims to evaluate quality of life and associated characteristics among epilepsy patients who visited North Shoa zone hospitals in Ethiopia.

**Methods:** An institution-based cross-sectional study design was conducted from April -May 2021 at North Shoa zone hospitals. A systematic random sampling technique was used to get a total number of 472 samples. Data on quality of life was assessed through interviews using the World Health Organization Quality of Life—Brief (WHOQOLBREF) Version. The collected data were coded, entered into Epi Data 3.1, and analyzed by using SPSS version 25. Simple and multiple linear regression analysis models were fitted and the unstandardized β coefficient at 95% confidence interval was employed. The statistical significance was accepted at *p*-value <0.05.

**Results:** From a total of 472 participants the response rate was 98%. The mean score of quality of life was 57.2±12.3. Age (β=5, 95% CI: 2.301, 7.699), marital status (β=-6.914, 95%CI: -8.867, - 4.961),seizure frequency (β=-.2.307, 95%CI: -4.795, .020), Anti-epileptic drug non-adherence (β=-.11.016, 95%CI: -13.642, -8.389), anxiety (β-4.062, 95%CI: (−6.503, -1.621), poor social support (β=-6.220, 95%CI: (−8.422, -4.017) and moderate social support (β=-5.58, 95%CI: -7.792, -3.368) were significantly associated with quality of life.

**Conclusion:** The mean quality of life of people living with epilepsy in this study was low. Age, marital status, seizure frequency, concomitant anxiety, antiepileptic drug non-adherence, number of anti-epileptic drugs/day, moderate and poor social support were all found to be predictors of quality of life in this study. As a suggestion, the patient treatment plan should include a quality of life assessment that addresses psychosocial concerns; professional counseling on how to cope with psychological, environmental, and social difficulties should be increased.

## 1. Introduction

Quality of life can be defined as “individuals’ perceptions of their position in life in the context of the culture and value systems in which they live and, in relation to their goals, expectations, standards and concerns (1). It is a broad term that incorporates the physical health, psychological condition, degree of freedom, social relationships, personal values and their relationships to important environmental characteristics in a complex manner (2). An ideal health assessment, therefore, would include a measure of the person’s physical health, a measure of physical, social and psychological functioning, and a measure of quality of life. To devise a measure of quality of life that is both reliable and valid, a broad range of potentially independent domains covering all important aspects of quality of life is necessary (3).

International League against Epilepsy (ILAE) defines epilepsy as a disease of the brain defined by any of the following conditions: (i) At least two unprovoked (or reflex) seizures occurring > 24 hrs. Apart; (ii) one unprovoked (or reflex) seizure and a probability of further seizures similar to the general recurrence risk (at least 60%) after two unprovoked seizures, occurring over the next 10 years; or (iii) diagnosis of an epilepsy syndrome (4).

The disease has significant effects on social, cognitive, psychological, and physical components of life as well as quality of life of the patients. The effect of epilepsy on quality of life is triggered by several factors including the use of antiepileptic medications frequency of seizure duration on anti-epileptic drugs (AED) (5). Epilepsy is a devastating disorder that affects patients’ quality of life, regardless of use of antiepileptic drugs (AEDs) (6).

Epilepsy is the most common neurological disease that affects around 50 million people of all ages globally. All over the world, epileptic patients are the focus of human rights violations and discrimination. The stigma of epilepsy can discourage people from seeking health care and has a negative impact on quality of life and social enclosure (7).

Epilepsy accounts for over 13 million disability-adjusted life years (DALYs) and is accountable for more than 0.5% of the global burden of disease (GBD) (7). Psychiatric and Physical comorbidities in people with epilepsy are linked with increased health care needs, poorer health outcomes, greater social exclusion and decreased quality of life. Anxiety (20%) and depression (23%), respectively, are the most common mental comorbidities (8).

The impact of epilepsy on quality of life can be significant with extensive and life-long consequences; the key areas where quality of life is influenced by epilepsy include Education, Employment, independence and social isolation (8).

The study at Gonder university hospital revealed that marital status did not significantly affects quality of life (9) but other studies revealed that marital status significantly affects quality of life (10-13).

According to many researches on epileptic patients’ quality of life, clinical and personal traits are proximal or direct determinants, whereas socio demographic factors are distant factors that influence epileptic patients’ quality of life (9-11, 13, 14).

Therefore, the aim of this study was to assess quality of life and associated factors among patients with epilepsy, and the results of this study will add an input to policy makers to scale up a public health program for designing and implementing strategies to improve quality of life.

## 2. Methods and Materials

### 2.1 Study area and period

The study was conducted at North shoa zone hospitals, Amhara regional state from April 01/4/2021-May 30/05/2021. North shoa zone is one of 11 Zones in the Ethiopian Amhara Region. The Zone is bordered on the south and the west by the Oromia Region, on the north by South Wollo, on the northeast by the Oromia Zone, and on the east by the Afar Region. Based on the 2018 demographic and health data, the zone has a total population of 1,692,555(15). The zone has 10 public, two private hospitals, 99 health centers, 391 health post,and 164 private clinics which provide different health services for the urban community and, people coming from different neighboring regions.

### 2.2 Study design

Institutional based cross-sectional study design was employed.

### 2.3 Populations

#### 2.3.1 Source population

All epileptic patients on-treatment follow up at hospitals of north shoa zone.

#### 2.3.2 Study population

All epileptic patients attending at the selected hospitals of north shoa zone during the study period.

#### 2.3.3 Study unit

Each individuals who participate in the study at the selected hospitals.

### 2.4 Inclusion and Exclusion criteria

#### 2.4.1 Inclusion criteria

Patients with epilepsy age 18 years and above who are on antiepileptic medication at least for six months were included in the study.

#### 2.4.2 Exclusion criteria

Patients who are mentally unstable and critically ill patients were excluded.

### 2.5 Sample Size Calculation

Sample size is determined using a single population mean formula with the assumptions of 61±11.6 mean and standard deviation of the overall quality of life of a study conducted in Ethiopia (16)

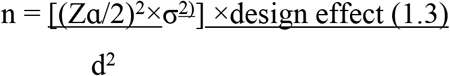

Since the source population was < 10, 000, correction formula was used and by considering 10% non-response rate, the final calculated sample size was **472**.

### 2.6 Sampling technique

12 hospitals in North Shoa zone were stratified based on the service they provide (comprehensive specialized, general, and primary hospitals). Simple random selection was applied for the strata that contain more than one hospital based on 30% coverage to secure representativeness. The required sample size was taken proportionally to the size of the selected hospitals. To get the individual sample units (participants) at the selected hospitals systematic random sampling was conducted by using the total number of epileptic individuals who have an appointment during the study period and number of samples required in each selected hospital.

After getting the sampling fraction in the selected hospital a simple random method/lottery method was used among the first “k” units to get the first participant.

### 2.7 Data collection methods

Data was collected using data abstraction format and the standardized WHOQOL-BRIEF questionnaire(1). Data regarding drug adherence, perceived stigma, self-esteem, social support, sleep quality and substance use was collected by using questionnaire adopted from different tools.

### 2.8 Data collection instruments

WHOQOL-BREF contains 26 items and a sound, cross-culturally valid assessment of QOL, consisting of four domains (6): physical health (7 items), psychological health (6 items), social relationships (3 items), and environmental health (8 items); it also contains the first two questions on the general perception of life and health and Each individual item of the WHOQOL-BREF were scored from 1(very dissatisfied/very poor) to 5 (very satisfied/very good).

Drug adherence: was measured by 10 items medication adherence rating scale; MARS (17).

HADS was a 14-item questionnaire, commonly used to screen for symptoms of anxiety and depression. The 14-items can be separated into two 7-item subscales for anxiety and depression (18).

Self-esteem: was measured by 10 items of Rosenberg self-esteem scale (19)

Perceived stigma: was measured by revised version of the Epilepsy Stigma Scale (rESS) with 3 items questionnaire on a three-point rating scale (0 = not at all, 1 = yes, maybe, 2 = yes, probably, 3 = yes, definitely) (20)

Social support: was measured by 14 items of Oslo social support scale (OSSS-3) (21).

Sleep quality: was measured by piths burgh sleep quality index 19 items of questionnaire (22).

### 2.9 Dependent and Independent variables

#### 2.9.1 Dependent Variables

➢ Quality of Life of epileptic patients.

#### 2.9.2 Independent Variables

➢ ***Socio-demographic factors:*** Sex, Age, Religion. Educational status, marital status, Occupation, residence.
➢ ***Clinical Factors:*** seizure type, seizure frequency, number of AED /day, Duration of epilepsy in yrs., history of comorbid disease, family history of epilepsy, duration on-treatment, current status of epilepsy,
➢ ***Psychosocial factors:*** Perceived self-esteem, perceived stigma, Compliance to self-care, social support, enough and regular sleep, Recreational activity, Regular physical activity, Ever substance use, Current substance use.

### 2.10. Operational and term definition

- ***Quality of life:-***the physical, psychological, social and environmental well-being of patients with epilepsy.
- Based on HAD scale classified anxiety &depression if the total score is (18):-

✓ **0-7 = Normal**: - patients who have no sign of depression and anxiety.
✓ **8-10 = Borderline abnormal (borderline case):-** patients who have at borderline or have risk developing depression and anxiety.
✓ **11-21= Abnormal (case):-** patients who had sign or case of depression and anxiety.

- ***Drug adherence***: based on medication adherence rating scale Patients who scored below 7 were considered as no adherent to their antiepileptic medications (17).
- ***Perceived self-esteem****:-* based on Rosenberg self-esteem scale Higher scores indicates higher self-esteem (19).
- ***Perceived stigma***: based on revised version of the Epilepsy Stigma Scale 0 represents no stigma, a score of 1–6 represents mild to moderate stigma, and a score of 7–9 represents high stigma.
- ***Social support***: based on Oslo social support scale individuals with total score 3-8 poor social support, 9-11 moderate social support, and 12-14 strong social support (21)
- ***Sleep quality***: **Poor sleep quality**: explained by a cut-off point of greater than or equal to 5 by using PSQI (23).
- ***Good sleep quality***: explained by a cut-off point of less than to 5 by using PSQI (23).
- ***Regular physical activity***: Considered if a person practiced any activity including walking for more than 30 minutes a day.
- ***Current substance use***: use of drugs or alcohol, cigarettes, illegal drugs, prescription drugs, inhalants at the moment.

### 2.11 Data quality assurance and control

The questionnaire and consent documents was first developed in English language, then translate into Amharic, local language, for data collection and finally retranslated back into English by another translator to check consistency. Face validity of the questionnaire was assessed and meanings of all items was checked before the actual data collection accordingly.

Items interpretability and understandability by the study participants were evaluated by pre-testing the questionnaire on epileptic patients (5% of the total sample size) having follow up at Arerti hospital and necessary correction was taken accordingly. Six data collectors who has psychiatric in nursing were training on overall project plan, using a rapport approach and on how to communicate with epileptic patients to elicit reliable data.

Supervision was also done on the spot by principal investigator and one supervisor. The collected data was checked for its completeness and clarity by the principal investigator and supervisor daily. Data clean up and cross-checking was done before analysis.

### 2.12 Data entry and analysis

After checking for its completeness, data was checked, cleaned, and entered into Epi data version 3.1 and transferred to SPSS version 25 for analysis. Descriptive statistics were expressed in frequency, percentage, mean and standardization to summarize socio demographic, clinical and personal characteristics and evaluate distribution of responses.

Linear regression model was used to assess the association between the dependent and different explanatory variables. Variables with p-value <0.2 were further analyzed by multiple linear regression and value < 0.05 take as level of significance.

The psychometric property WHO QOL-BREEF questionnaire was tested for its reliability by author’s Cranach’s alpha test value of >0.7 as accepted internal consistency.

### 2.13 Ethical consideration

The respondents’ rights and dignity was respected. Written consent was obtained from the study participant (for the minors especially for those <18yrs consent was obtained from their parents) to confirm willingness for participation after explaining the objective of the study. The respondents were notified that they have the right to refuse or terminate at any time of the interview. The information provided by each respondent was kept confidential throughout the research process.

## 3. Results

### 3.1 Socio-demographic characteristics

A total of 462 participants took part with a response rate of 98%. The mean age of the participants was 33.31 (±SD=13.7) years. Among the participants 297(64.3%) were males and 165(35.7) were females. Majority of the participants, 145(31.4%) were within the age group of 25-34yrs (**See table below**).

**Table 1.**
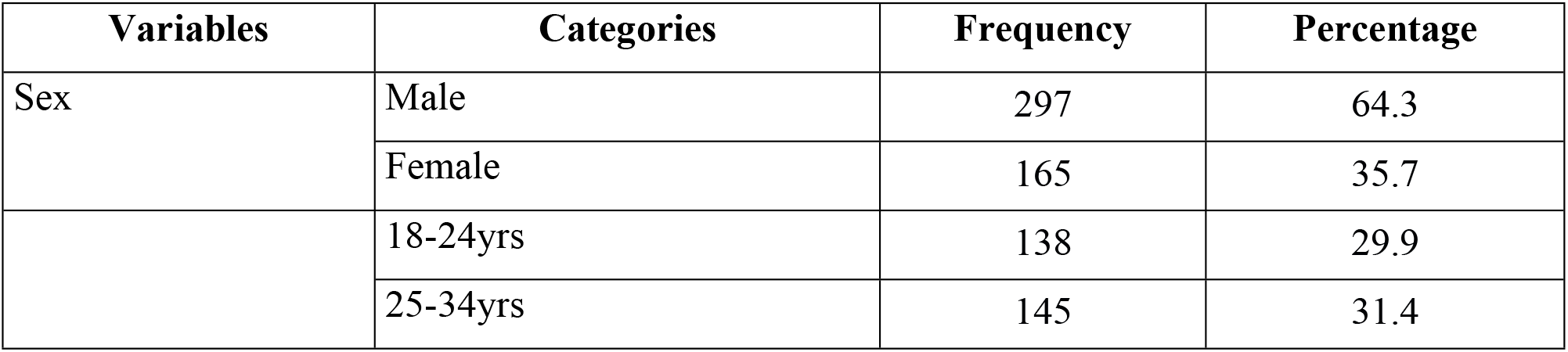
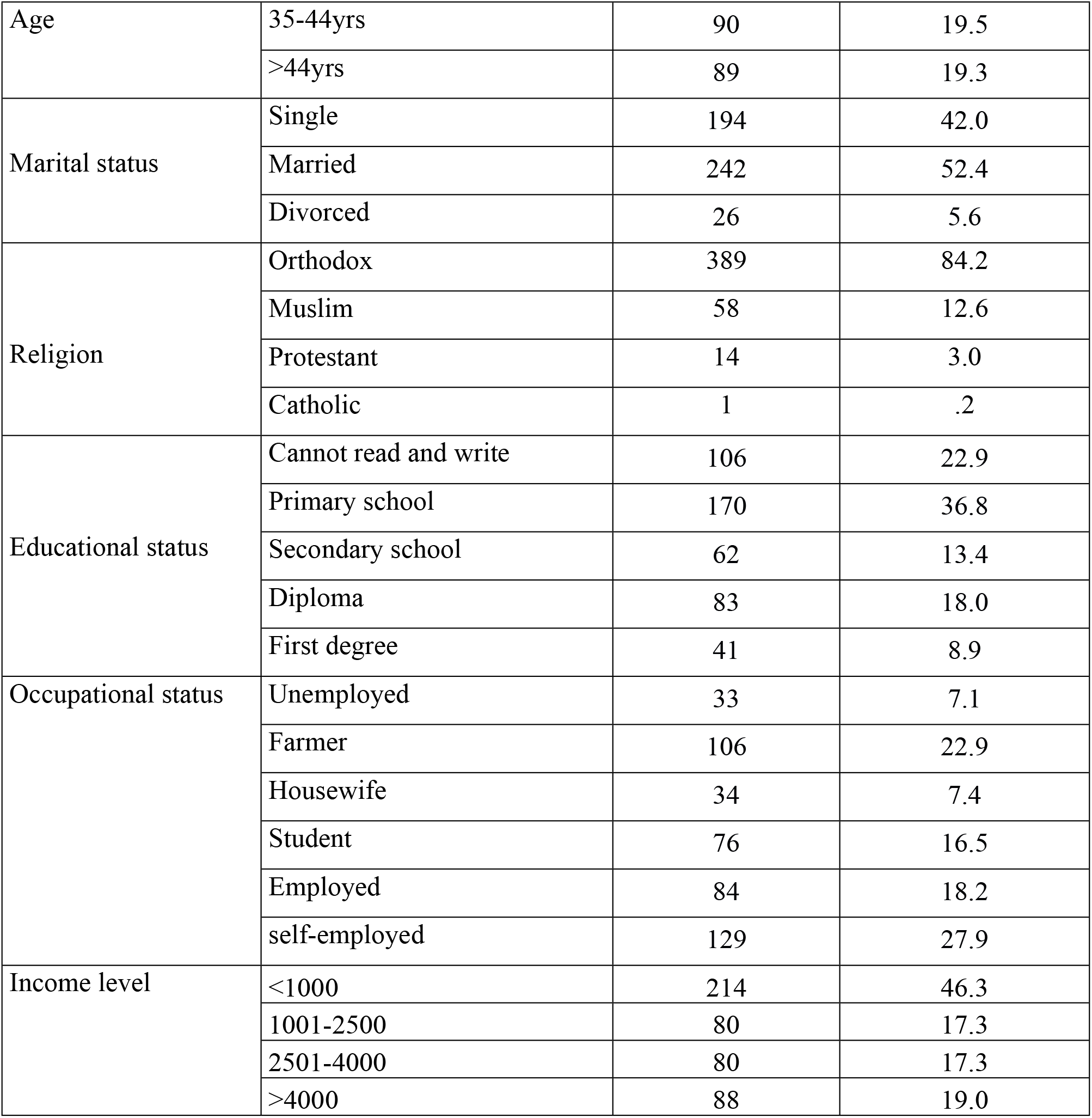
Socio demographic characteristics of epileptic patients on treatment follow up in North Shoa Administration, Amhara national state, Ethiopia, 2021 (N=462)

### 3.2 Clinical characteristics

The mean age for the onset of epilepsy was 22.68 (±SD=12.8) years. Regarding the duration of the illness, 187 (40.5%) had 11 and above years. The majority of the participants, 377(81.6%) were under mono therapy, and 220 (47.6%) had one or more seizure attack during last follow up. The mean of AED adherence was 7.2±0.82.(**See table below**).

**Table 2:** Clinical characteristics of epileptic patients on treatment follow up in North Shoa admiration, Amhara national state, Ethiopia, 2021 (N=462)

| Variables | Categories | Frequency | Percentage |
| --- | --- | --- | --- |
| Age at diagnosis | <=15yrs | 143 | 31.0 |
|  | 16-30yrs | 222 | 48.1 |
|  | 31-45yrs | 61 | 13.2 |
|  | >45yrs | 36 | 7.8 |
| Type of epilepsy | generalized onset | 397 | 85.9 |
|  | focal onset | 65 | 14.1 |
| Seizure frequency during last follow up | no seizure | 159 | 34.4 |
|  | 1-2 seizure | 220 | 47.6 |
|  | 3 or more seizure | 83 | 18.0 |
| Number of AED/day | Mono therapy | 387 | 83.8 |
|  | Poly therapy | 75 | 16.2 |
| Epilepsy duration | <=5yrs | 128 | 27.7 |
|  | 5-10yrs | 147 | 31.8 |
|  | >=11 yrs | 187 | 40.5 |
| Treatment duration | 1-5yrs | 190 | 41.1 |
|  | 6-10 | 148 | 32.0 |
|  | >10 | 124 | 26.8 |
| Current drug(s) | Phenobarbitone | 288 | 62.3 |
|  | Carbamazepine | 71 | 15.4 |
|  | Phenobarbitone and valproate | 20 | 4.3 |
|  | Phenobarbitone and carbamazepine | 42 | 9.1 |
|  | Valproate | 28 | 6.1 |
|  | Phenytoin | 9 | 1.9 |
|  | Others* | 4 | .9 |
| Drug adherence | Adherent | 408 | 88.3 |
|  | Non adherent | 54 | 11.7 |
| Comorbidity of anxiety | Yes | 67 | 14.5 |
|  | No | 395 | 85.5 |
| Comorbidity of depression | Yes | 59 | 12.8 |
|  | No | 402 | 87.0 |
| Other comorbid diagnosed disease<sup>a</sup> | Yes | 74 | 16.0 |
|  | No | 388 | 84.0 |
| Current status of epilepsy | No change | 36 | 7.8 |
|  | Improved | 404 | 87.4 |
|  | Deteriorated | 22 | 4.8 |

**Notes:** <sup>a</sup> =Diabetes mellitus, HIV/AIDS, hypertension
\* = clonazepam, lamotrigine

### 3.3 Psychosocial factors

Regarding personal and social characteristics of the participants, 236(51.1%) had perceived stigma; 212(45.9%) had poor social support. About 377(81.6%) had good Sleep quality and 49 (10.6%) of the participants were ever used substances for a non-medical purpose and the mean score of self-esteem of the participants were 20.1±3.7(**See table below**).

**Table 3:** Personal (individual and social) characteristics of epileptic patients on treatment follow up in North Shoa administration, Amhara national state, Ethiopia, 2021 (N=462)

| *Variables* | *Categories* | *Frequency* | *Percentage* |
| --- | --- | --- | --- |
| Perceived stigma | had no stigma | 236 | 51.1 |
|  | had stigma | 226 | 48.9 |
| Social support | Poor | 212 | 45.9 |
|  | Moderate | 142 | 30.7 |
|  | Strong | 108 | 23.4 |
| Self esteem |  | Mean =20.1±3.7 |  |
| Sleep quality | good sleep quality | 377 | 81.6 |
|  | poor sleep quality | 83 | 18.0 |
| Substance use | no substance use | 413 | 89.4 |
|  | substance user | 49 | 10.6 |

### 3.4 Mean Quality of Life Score of Participants

The overall mean (SD) score of quality of life among epilepsy patients was 57.2±12.3. Among four domains of WHO QOL-BREF, participants scored highest in the physical domain (62.43±6.5), while the lowest mean (SD) score was for environment domain (49.59 ±7.03)(**See table below**)

**Table 4:** Mean scores of Quality of Life scale of epileptic patients on treatment follow up in North Shoa administration, Amhara national state, Ethiopia, 2021 (N=462)

| Variables | Likert scale Mean<br>± SD | Formula for domain<br>mean* | Domain mean ± SD of 100 |
| --- | --- | --- | --- |
| Physical domain (7 items) | $3.5 \pm 0.93$ | $100 / (35 - 7) \times (24.48 - 7)$ | $62.43 \pm 6.5$ |
| Psychological domain (6 items) | $3.6 \pm 0.9$ | $100 / (30 - 6) \times (20.17 - 6)$ | $59.04 \pm 5.6$ |
| Social domain (3 items) | $3.29 \pm 0.78$ | $100 / (15 - 3) \times (9.88 - 3)$ | $57.3 \pm 2.34$ |
| Environmental (8 items) | $2.98 \pm 0.88$ | $100 / (5 - 1) \times (3.56 - 1)$ | $49.59 \pm 7.03$ |
| How would you rate your QOL | $3.56 \pm 0.727$ | $100 / (5 - 1) \times (3.56 - 1)$ | $64 \pm 0.727$ |
| How satisfied are you with your health | $3.49 \pm 0.784$ | $100 / (5 - 1) \times (3.49 - 1)$ | $62.25 \pm 0.784$ |
| Overall HRQOL | $3.29 \pm 0.8$ | $100 / (130 - 26) \times (85.45 - 26)$ | $57.2 \pm 12.31$ |
| *Domain mean transformed to 100 = $100 / (\text{maximum score} - \text{no of items}) \times (\text{sum of means} - \text{no of items})$ | | | |

### 3.5 Factors Associated with Quality of Life

A simple linear regression was performed in connection to a variety of variables that could potentially influence quality of life. Variables with *p*-value <0.20 during simple linear regression analysis were selected for further analysis in multiple linear regression analysis.

#### 3.5.1 Associated factors simple linear regression analysis result

Age, marital status, seizure frequency, number of antiepileptic drug, epilepsy duration, duration on treatment, anxiety and depression, current status of epilepsy, drug adherence, self-esteem, perceived social stigma, social support, comorbidity of other medical illness and sleep quality were factors associated with quality of life in simple linear regression analysis(**See table below**).

**Table 5:**
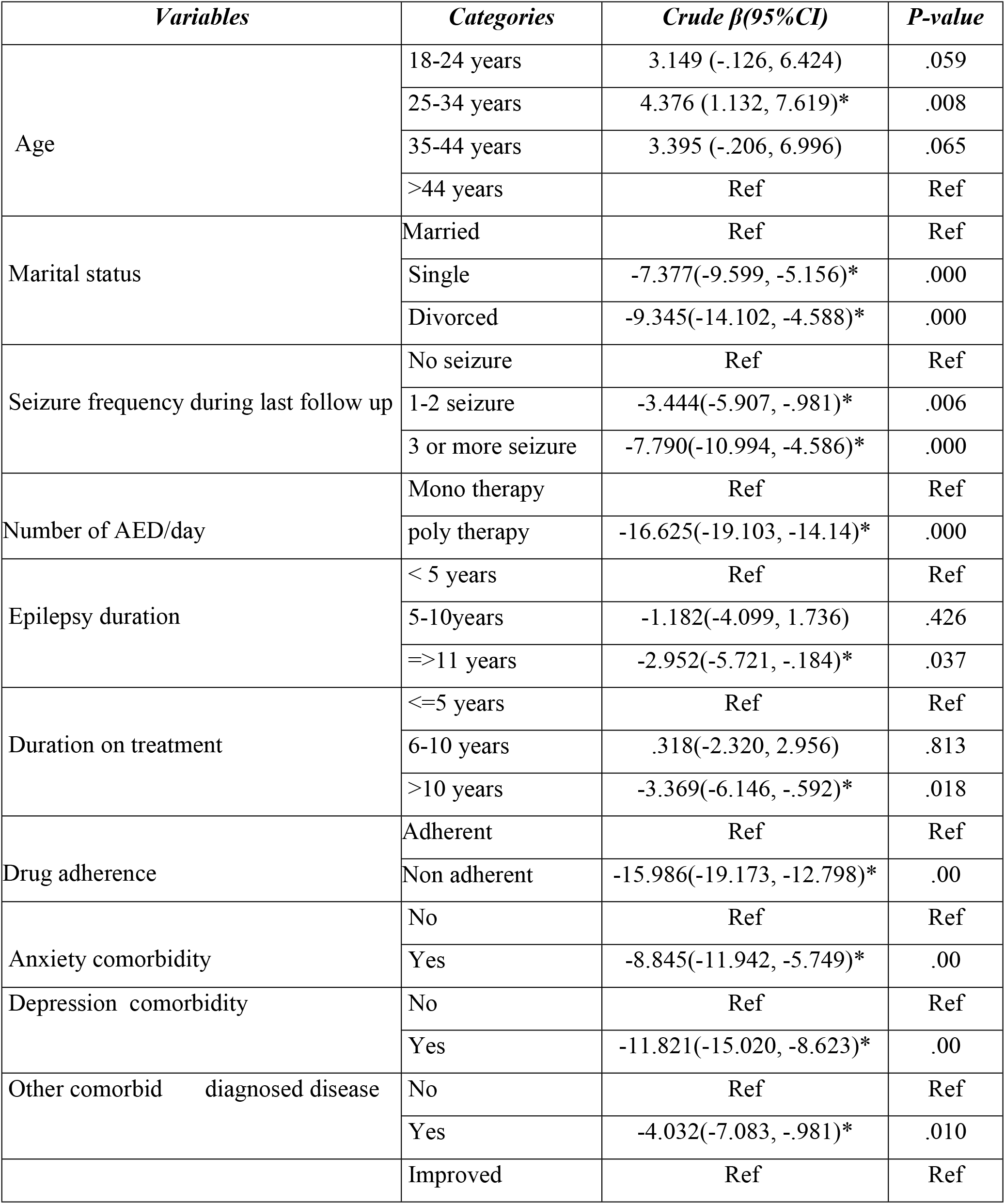
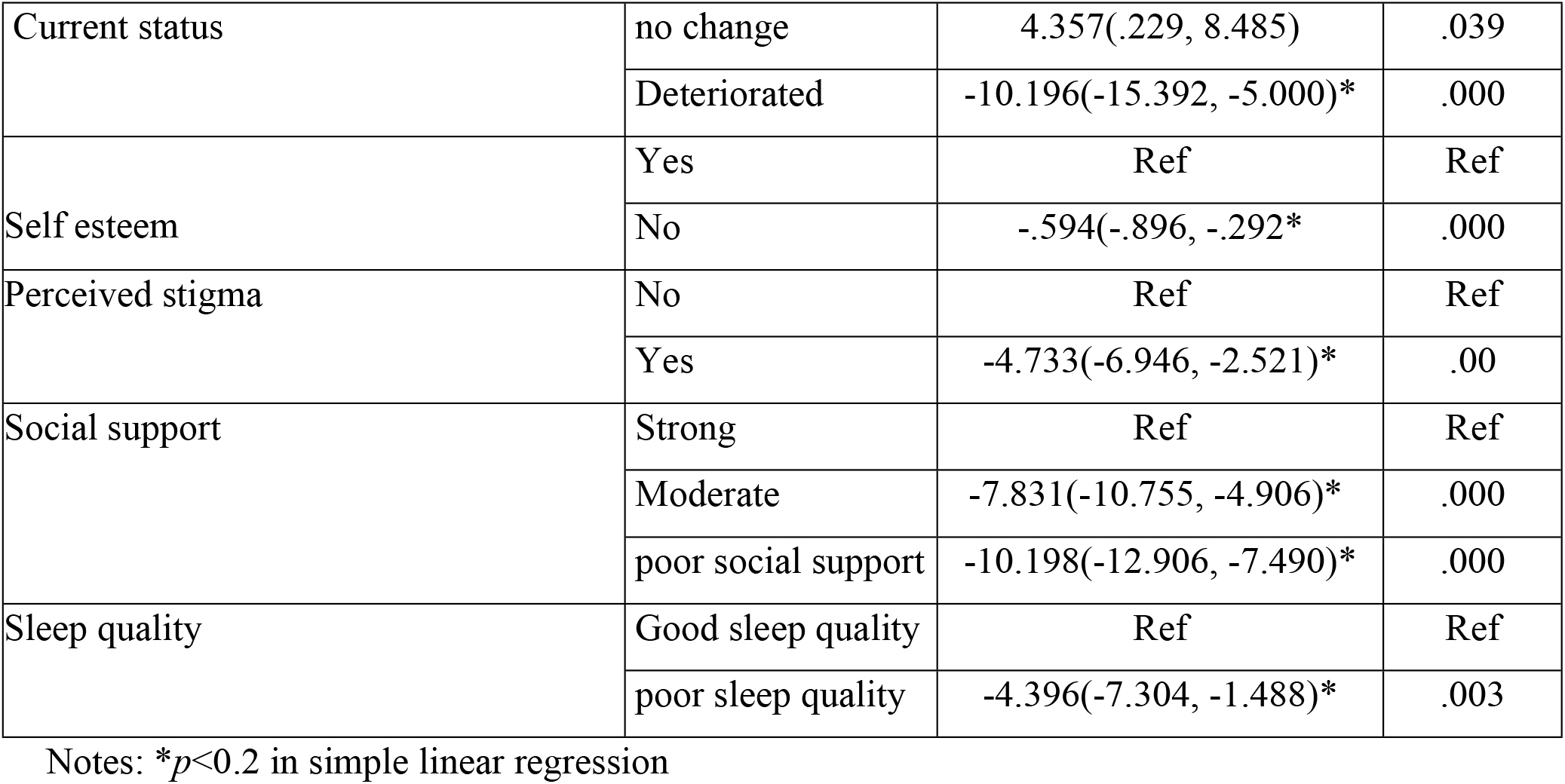
Simple linear regression analysis of factors associated quality of life of epileptic patients on treatment follow up in North Shoa administration, Amhara national state, Ethiopia, 2021

#### 3.5.2 Associated factors multiple linear regression analysis result

The results of multiple linear regression showed that age, marital status, seizure frequency, poly drug therapy, AED non-adherence, anxiety, and poor social support were found to be statistically significant at *p*-value <0.05.

A score in quality of life is increased by 5 at the age group 18-24(β=5, 95%CI: 2.301, 7.699) compared to the age group above 44 years. A score in quality of life decreased by 6.9 in single individuals compared to married individuals (β=-6.914, 95%CI: -8.867, -4.961).

A score in quality of life decreased by 2.3 in every 3 or more seizure attacks during last follow up; (β=-2.307, 95%CI: -4.795, .020). A score in quality of life decreased by 12.7 in every unit increase of poly-therapy score (β=-12.661, 95%CI: -15.030, -10.293). Quality of life of epileptic patients decreased by 4.06 in every unit increase in a score of anxiety symptoms (β=-4.062, 95%CI: - 6.503,-1.621)(**See table below**).

**Table 6:** Multiple Linear Regression of Quality of Life of epileptic patients on treatment follow up in North Shoa administration, Amhara national state, Ethiopia, 2021 (N=462)

| *Variables* | *Categories* | *Unstandardized $\beta$* | *standardized $\beta$* | *95%CI* |
| --- | --- | --- | --- | --- |
|  | 18-24 years | 5.000 | .186 | (2.301, 7.699)* |
| Age | 25-34 years | 3.298 | .124 | (.901, 5.695) |
|  | 35-44 years | .856 | .028 | (-1.697, 3.409) |
|  | >44 years | Ref | Ref | Ref |
| Marital status | Married | Ref | Ref | Ref |
|  | Single | -6.914 | -.277 | (-8.867, -4.961)* |
|  | Divorced | -2.850 | -.053 | (-6.429, .729) |
| Seizure frequency during last follow up | No seizure | Ref | Ref | Ref |
|  | 1-2 seizure | .298 | .012 | (-1.571, 2.168) |
|  | 3 or more seizure | -2.387 | -.075 | (-4.795, .020)* |
| Number of AED/day | Mono therapy | Ref | Ref | Ref |
|  | poly therapy | -12.661 | -.399 | (-15.030, -10.293)* |
| Epilepsy duration | < 5 years | Ref | Ref | Ref |
|  | 5-10years | .497 | .019 | (-2.120, 3.115) |
|  | =>11 years | -.988 | -.039 | (-3.883, 1.908) |
| Duration on treatment | <=5 years | Ref | Ref | Ref |
|  | 6-10 years | 1.012 | .038 | (-1.397, 3.421) |
|  | >10 years | -.838 | -.030 | (-3.697, 2.021) |
| Drug adherence | Adherent | Ref | Ref | Ref |
|  | Non adherent | -11.016 | -.288 | (-13.642, -8.389)* |
| Anxiety comorbidity | No | Ref | Ref | Ref |
|  | Yes | -4.062 | -.116 | (-6.503, -1.621)* |
| Depression comorbidity | No | Ref | Ref | Ref |
|  | Yes | -2.163 | -.059 | (-5.018, .692) |
| Other comorbid diagnosed disease | No | Ref | Ref | Ref |
|  | Yes | -.633 | -.019 | -2.854, 1.589) |
| Current status | Improved | Ref | Ref | Ref |
|  | no change | -.309 | -.007 | (-3.305, 2.687) |
|  | Deteriorated | -.222 | -.004 | (-4.248, 3.803) |
| Self esteem | Yes | Ref | Ref | Ref |
|  | No | .195 | .058 | (-.049, .440) |
| Perceived stigma | No | Ref | Ref | Ref |
|  | Yes | -1.114 | -.045 | (-2.802, .575) |
| Social support | Strong | Ref | Ref | Ref |
|  | Moderate | -5.580 | -.209 | -7.792, -3.368)* |
|  | poor social support | -6.220 | -.252 | -8.422, -4.017)* |
| Sleep quality | Good sleep quality | Ref | Ref | Ref |
|  | poor sleep quality | -.214 | -.007 | (-2.371, 1.943) |

Notes: \* $p < 0.05$ in multiple linear regression and statically significant

## 4. Discussion

This study tried to assess quality of life and associated factors among patients with epilepsy. According to this study finding, the mean quality of life score among participants was 57.2 (±SD=12.3). This result was in line with 61.49 mean score of Indian (24),58 mean score of Ugandan(6), 60.47 mean score of Wollega zone (10),58 mean score of South Wollo and 60.14 mean score of Amanuel mental specialized hospital (16) studies.

In this study, the mean quality of life score in epileptic patients is higher, compared to a study conducted in Kenya (mean score of 49.90), and Bhutan (mean score of 48.9) (25, 26). This difference could be due to sample size, clinical characteristics of the participants, cultural differences, or the study setting. When compared to the Kenyan and Bhutanese samples, this study had a larger sample size.

In the Kenyan study, the participants use of poly AED therapy was large (54%) compared with this study (16.2%). Different literature (27, 28) has shown poly therapy receiving patients had lower mean quality of life. More than half (69%) of study participants in Poland had medical comorbidity compared with 16% in this study. This high comorbidity of medical illness might contribute to low mean quality of life.

This result, however, was lower than the average score of other countries like 74.9 in India, 70.64 in Serbia, 68.9 in Malaysia, 82 in Canada, and 66.0 in the UK (41, 22, 24, 41-42). The availability of standard medical care, sociocultural beliefs, and the patient’s lifestyle, as contrasted to this study, could be the explanation for this improved quality of life.

About domains of quality of life, the environmental domain had low quality of life (mean score of 57.78). This result was supported by Indian and Addis Ababa studies, where the environmental domain of patients with epilepsy was mostly affected (42, 26).

In contrast, results from Ghana showed that the social domain score was the lowest mean score (29). The disparity might be attributed to differences in belief, culture, and lifestyle variables, all of which have an impact on quality of life measurements. Because quality of life is a subjective notion, people from various cultures may interpret it differently.

When compared to those who were married, the study found that being single was connected with a lower quality of life. This result was supported by other studies in India and Wollega zone (24, 30) in Ethiopia. One probable reason for this outcome is that single people will take on more responsibility for their everyday lives on their own, resulting in a weaker psychosocial area of life.

The study’s findings also demonstrated that getting older has a detrimental impact on quality of life. Studies in India and Australia backed up this conclusion (24, 31). The reason for this could be because epilepsy in older people results in lower psychosocial functioning when compared to healthy controls (31). Poly therapy was negatively associated with quality of life; this result was supported by the study in Nigeria (28). The possible explanation might be related to the side effect of AED.

Seizure frequency was significantly associated with quality of life. This finding was consistent with other studies (32-36). The most likely explanation is that frequent seizures are linked to excessive fear, a sense of stigmatization, and a reduction in physical activity. This could lead to a reduction in physical activity as well as psychological issues.

According to this study Patient with comorbid anxiety scored lower on the quality of life scale, Studies in Thailand, United Kingdom, Wollega zone and Amanuel specialized mental hospital in Addis Ababa supported this finding (32, 37). Patients with epilepsy who experience frequent seizures showed increased anxiety symptoms (38) and such a mental health issue is not addressed, It would have a negative consequence to quality of life of patients.

The result of this study showed that AED Non-adherence had negatively associated with quality of life. This finding was supported by studies in UK (32) and US (31). Patients who were non-adherent were more likely to have had severe life consequences as a result of their seizures (39).

The possible explanation might be Non-adherence to an antiepileptic drug (AED) may result in poor clinical outcomes and increased seizure frequency, resulting in a lower quality of life.

Poor social support was associated with a lower quality of life. The experience of being liked and wanted could have contributed to creating a supportive environment that helped people cope with their disease. This finding was supported by another study in Switzerland (40)

## 5. Conclusion

The mean quality of life of people living with epilepsy in this study was low. The finding from this study also indicated that age, marital status, frequent seizure, comorbid anxiety, antiepileptic drug no adherence, poly therapy, moderate and poor social support were the predictors of quality of life.

## 6. Recommendations

### To Debre Birhan zonal health department

- Incorporating quality of life assessment tool to patient assessment and treatment plan formats.
- Creating agenda on how to improve the quality of life of epileptic patients and reduce associated factors.
- Monitoring and evaluation of each health care units how to handle epileptic patients.

### To health care professionals

- Increase professional counseling on how to cope with psychological, environmental, and social difficulties.
- Implementing therapies that focus on early detection of concomitant psychiatric diseases in patients with epilepsy, such as depression and anxiety.

### To Debre Birhan university

- Giving training for health care professionals specially focusing on the four domains of quality of life.

## Data Availability

values behind the means, standard deviations and other measures are reported as well points are extracted from images for analysis.

## Authors’ contributions

EG-performed data collections and analyzed the data. EG, GE, BT, NA & AE wrote the manuscript. All authors read and approved the final manuscript.

## Acknowledgements

We would like to acknowledge Debre Birhan University and CBE office for financial and technical support. We also would like to express our heartfelt thanks to our advisor Mr. **Akine Eshete** (BSc, MPH and Assistance professor of public health) and **Mr. Nigus Alemnewu** (BSc, MSc) for their genuine and valuable comment given for us.

## References

1. Organization WH. WHOQOL-BREF: introduction, administration, scoring and generic version of the assessment: field trial version, December 1996. World Health Organization, 1996.

2. Organization WH. Programme on mental health: WHOQOL user manual. World Health Organization, 1998.

3. Group TW. The World Health Organization quality of life assessment (WHOQOL): development and general psychometric properties. Social science & medicine. 1998;46(12):1569–85.

4. Fisher RS, Acevedo C, Arzimanoglou A, Bogacz A, Cross JH, Elger CE, et al. ILAE official report: a practical clinical definition of epilepsy. Epilepsia. 2014;55(4):475–82.

5. Gebre AK, Haylay A. Sociodemographic, clinical variables, and quality of life in patients with epilepsy in Mekelle City, Northern Ethiopia. Behavioural Neurology. 2018;2018.

6. Nabukenya AM, Matovu JK, Wabwire-Mangen F, Wanyenze RK, Makumbi F. Health-related quality of life in epilepsy patients receiving anti-epileptic drugs at National Referral Hospitals in Uganda: a cross-sectional study. Health and quality of life outcomes. 2014;12(1):49.

7. Organization WH. Epilepsy: a public health imperative: World Health Organization; 2019.

8. UCB. UCB. Epilepsy and Quality of Life: Fact Sheet. 2008. 2008.

9. Muche EA, Ayalew MB, Abdela OA. Assessment of Quality of Life of Epileptic Patients in Ethiopia. International Journal of Chronic Diseases. 2020;2020.

10. Abadiga M, Mosisa G, Amente T, Oluma A. Health-related quality of life and associated factors among epileptic patients on treatment follow up at public hospitals of Wollega zones, Ethiopia, 2018. BMC research notes. 2019;12(1):679.

11. Stotaw AS, Beyene DA, Hasen SL, Ali YS. Health Related Quality of Life and Associated Factors among Epileptic Patients at Dessie Referral Hospital, Amhara Regional State, North East Ethiopia, 2020, A Cross Sectional Study.. 2020.

12. Tefera GM, Megersa WA, Gadisa DA. Health-related quality of life and its determinants among ambulatory patients with epilepsy at Ambo General Hospital, Ethiopia: Using WHOQOL-BREF. PloS one. 2020;15(1):e0227858.

13. Tegegne MT, Muluneh NY, Wochamo TT, Awoke AA, Mossie TB, Yesigat MA. Assessment of quality of life and associated factors among people with epilepsy attending at Amanuel Mental Specialized Hospital, Addis Ababa, Ethiopia. Science Journal of Public Health. 2014;2(5):378–83.

14. Alsaadi T, Kassie S, El Hammasi K, Shahrour TM, Shakra M, Turkawi L, et al. Potential factors impacting health-related quality of life among patients with epilepsy: results from the United Arab Emirates. Seizure. 2017;53:13–7.

15. Adugna A. Demography and Health 2018 [cited 2018 december 16].

16. Mesafint G, Shumet S, Habtamu Y, Fanta T, Molla G. Quality of Life and Associated Factors Among Patients with Epilepsy Attending Outpatient Department of Saint Amanuel Mental Specialized Hospital, Addis Ababa, Ethiopia, 2019. Journal of Multidisciplinary Healthcare. 2020;13:2021.

17. Thompson K, Kulkarni J, Sergejew A. Reliability and validity of a new Medication Adherence Rating Scale (MARS) for the psychoses. Schizophrenia research. 2000;42(3):241–7.

18. HADS). Hospital Anxiety and Depression Scale (HADS) [cited 2020 December 16/2020].

19. Rosenberg M. Rosenberg self-esteem scale (RSE). Acceptance and commitment therapy Measures package. 1965;61(52):18.

20. Taylor J, Baker GA, Jacoby A. Levels of epilepsy stigma in an incident population and associated factors. Epilepsy & Behavior. 2011;21(3):255–60.

21. Van Lente E, Barry MM, Molcho M, Morgan K, Watson D, Harrington J, et al. Measuring population mental health and social well-being. International journal of public health. 2012;57(2):421–30.

22. Buysse DJ, Reynolds III CF, Monk TH, Berman SR, Kupfer DJ. The Pittsburgh Sleep Quality Index: a new instrument for psychiatric practice and research. Psychiatry research. 1989;28(2):193–213.

23. Adem K, Kassew T, Birhanu A, Abate A. Sleep Quality and Associated Factors among Peoples with Epilepsy Who Have a Follow-Up at Amanuel Mental Specialized Hospital, Addis Ababa, Ethiopia, 2019: An Institutional Based Cross-Sectional Study. Psychiatry Journal. 2020;2020.

24. Rakesh P, Ramesh R, Rachel P, Chanda R, Satish N, Mohan V. Quality of life among people with epilepsy: a cross-sectional study from rural southern India. Natl Med J India. 2012;25(25):261–4.

25. Kinyanjui DW, Kathuku DM, Mburu JM. Quality of life among patients living with epilepsy attending the neurology clinic at Kenyatta National Hospital, Nairobi, Kenya: a comparative study. Health and quality of life outcomes. 2013;11(1):1–9.

26. Saadi A, Patenaude B, Nirola DK, Deki S, Tshering L, Clark S, et al. Quality of life in epilepsy in Bhutan. Seizure. 2016;39:44–8.

27. Staniszewska A, Kurkowska-Jastrzębska I, Tarchalska-Kryńska B. Quality of life in patients with epilepsy. Journal of Public Health, Nursing and Medical Rescue. 2015;157(2015_3):20–6.

28. Mosaku KS, Fatoye FO, Komolafe M, Lawal M, Ola BA. Quality of life and associated factors among adults with epilepsy in Nigeria. The International Journal of Psychiatry in Medicine. 2006;36(4):469–81.

29. Ankrah D. Pharmaceutical policies and access to medicines: a hospital-pharmacy perspective from Ghana: Utrecht University; 2017.

30. Abadiga M, Mosisa G, Amente T, Oluma A. Health-related quality of life and associated factors among epileptic patients on treatment follow up at public hospitals of Wollega zones, Ethiopia, 2018. BMC research notes. 2019;12(1):1–7.

31. McLaughlin DP, Pachana NA, Mcfarland K. Stigma, seizure frequency and quality of life: the impact of epilepsy in late adulthood. Seizure. 2008;17(3):281–7.

32. Ridsdale L, Wojewodka G, Robinson E, Landau S, Noble A, Taylor S, et al. Characteristics associated with quality of life among people with drug-resistant epilepsy. Journal of neurology. 2017;264(6):1174–84.

33. Norsa’adah B, Zainab J, Knight A. The quality of life of people with epilepsy at a tertiary referral centre in Malaysia. Health and quality of life outcomes. 2013;11(1):1–6.

34. Anu M, Suresh K, Basavanna P. A cross-sectional study of quality of life among subjects with epilepsy attending a tertiary care hospital. Journal of clinical and diagnostic research: JCDR. 2016;10(12):OC13.

35. Velez FF, Bond TC, Anastassopoulos KP, Wang X, Sousa R, Blum D, et al. Impact of seizure frequency reduction on health-related quality of life among clinical trial subjects with refractory partial-onset seizures: a pooled analysis of phase III clinical trials of eslicarbazepine acetate. Epilepsy & Behavior. 2017;68:203–7.

36. Piperidou C, Karlovasitou A, Triantafyllou N, Dimitrakoudi E, Terzoudi A, Mavraki E, et al. Association of demographic, clinical and treatment variables with quality of life of patients with epilepsy in Greece. Quality of Life Research. 2008;17(7):987–96.

37. Phabphal K, Geater A, Limapichart K, Satirapunya P, Setthawatcharawanich S. Quality of life in epileptic patients in southern Thailand. Medical journal of the Medical Association of Thailand. 2009;92(6):762.

38. Beyenburg S, Mitchell AJ, Schmidt D, Elger CE, Reuber M. Anxiety in patients with epilepsy: systematic review and suggestions for clinical management. Epilepsy & Behavior. 2005;7(2):161–71.

39. Hovinga CA, Asato MR, Manjunath R, Wheless JW, Phelps SJ, Sheth RD, et al. Association of non-adherence to antiepileptic drugs and seizures, quality of life, and productivity: survey of patients with epilepsy and physicians. Epilepsy & Behavior. 2008;13(2):316–22.

40. Mahrer-Imhof R, Jaggi S, Bonomo A, Hediger H, Eggenschwiler P, Krämer G, et al. Quality of life in adult patients with epilepsy and their family members. Seizure. 2013;22(2):128

